# Gaps in the Type 1 Diabetes Mellitus care cascade: a national perspective using South Africa’s National Health Laboratory Service (NHLS) database

**DOI:** 10.1101/2025.09.26.25336712

**Authors:** Alana T Brennan, Evelyn Lauren, Jacob Bor, Jaya A George, Siyabonga Khoza, Koleka Mlisana, Emma M Kileel, Sydney Rosen, Frederick Raal, Patricia Hibberd, Matthew P Fox, Nigel J Crowther

**Affiliations:** Department of Global Health, Boston University School of Public Health, Boston, MA, USA; Health Economics and Epidemiology Research Office, University of the Witwatersrand, Johannesburg, South Africa; Department of Epidemiology, Boston University School of Public Health, Boston, MA, USA; Department of Biostatistics, Boston University School of Public Health, Boston, MA, USA; Wits Diagnostic Innovation Hub, University of the Witwatersrand, Johannesburg, South Africa, Johannesburg, South Africa; National Health Laboratory Service, South Africa; Department of Chemical Pathology, University of the Witwatersrand, Faculty of Health Sciences, Johannesburg, South Africa; Division of Endocrinology and Metabolism, Department of Internal Medicine, Faculty of Health Sciences, University of the Witwatersrand, Johannesburg, South Africa

**Keywords:** type 1 diabetes mellitus, HIV/AIDS, tuberculosis, South Africa, care cascade

## Abstract

**Objective:** Type 1 diabetes mellitus (T1DM) requires lifelong management, yet access to insulin, specialized care, and education is limited in low- and middle-income countries (LMICs). While cascade-of-care analyses are well documented for type 2 diabetes (T2DM), to our knowledge no population-level estimates of the T1DM care cascade exist from LMICs. We therefore evaluated the T1DM care cascade nationally in South Africa and compared outcomes between youth living with HIV (YLWH) and youth living without HIV (YLWOH).

**Research Design and Methods:** We analyzed South Africa’s National Health Laboratory Service (NHLS) data for patients <20 years with a first glycated hemoglobin A1c (HbA1c) or plasma glucose [fasting (FPG), random (RPG)] between April 2004 and March 2015. Laboratory-diagnosed T1DM was defined as HbA1c ≥6.5%, FPG ≥7.0 mmol/L, or RPG ≥11.1 mmol/L. Cascade stages over 24 months were remaining in care and achieving glycemic control (HbA1c <7.0%, FPG <8.0 mmol/L, or RPG <10.0 mmol/L).

**Results:** Of 256,449 youth tested for diabetes, 12.1% met criteria for laboratory-diagnosed T1DM. Among these, 15.9% remained in care and 7.2% achieved glycemic control by 24 months. Retention was similar between YLWH and YLWOH (16.8% vs. 15.8%), but glycemic control was higher among YLWH (72.5% vs. 43.4%).

**Conclusions:** Four of five South African youth with T1DM lacked consistent care, and fewer than 10% achieved glycemic control within two years. Higher control rates among YLWH suggest lessons from HIV care models may strengthen T1DM services. These findings provide the first national estimates of the T1DM care cascade from sub-Saharan Africa and highlight the urgent need for targeted strategies to improve outcomes.

## Background

Between 2011 and 2021, sub-Saharan Africa recorded a five-fold increase in the prevalence of type 1 diabetes mellitus (T1DM) among children and adolescents.^1^ T1DM is a chronic autoimmune disorder resulting in the destruction of insulin-producing beta cells in the pancreas, leading to insulin deficiency and necessitating lifelong insulin therapy.^2^ It is one of the most common chronic diseases in childhood, arising primarily from genetic factors and environmental triggers including viral infections, dietary shifts, and environmental toxins.^3^ Poor healthcare infrastructure can lead to reduced access to diagnostic tools, inadequate patient education, and insufficient supplies of insulin and glucose monitors, making T1DM management in low- and middle-income settings particularly challenging and often increasing mortality rates.^4^

Research indicates that care for adults with type 2 diabetes (T2DM) in low- and middle-income countries (LMICs) often falls short of clinical guideline standards.^5-7^ Across the diabetes care continuum—from diagnosis to long-term management—services are frequently fragmented, contributing to inadequate care and poor health outcomes.^5-7^ Individuals with T1DM may face similar barriers; however, evidence on their care experiences remains limited. Given the distinct clinical and management needs of people with T1DM compared to those with T2DM, it is critical to identify the specific gaps in the T1DM care continuum to inform strategies for improvement.

South Africa is estimated to have approximately 4,500 people living with T1DM, ∼0.1% of the country’s more than 4 million people with diabetes overall; however, the unique clinical needs and lifelong insulin dependence of individuals with T1DM demand distinct attention within the health system.^8^ At the same time, South Africa has developed one of the world’s largest and most sophisticated public-sector infrastructures for HIV care, with nationwide systems for laboratory monitoring, patient tracking, and chronic disease management.^9^ This platform may influence retention and outcomes for individuals with T1DM, especially given the similar need for lifelong engagement in care. Understanding how HIV status shapes the T1DM care continuum offers important insight into whether existing infrastructure designed for people living with HIV (PLWH) can also be leveraged to support those with T1DM.

To evaluate the T1DM care continuum in South Africa’s public-sector facilities, we analyzed data from the National Health Laboratory Service (NHLS) database spanning 2004–2017. Our objectives were to determine the proportion of patients undergoing diabetes-related laboratory testing (glycated hemoglobin [HbA1c] or random/fasting plasma glucose) who were diagnosed with T1DM, remained in care, and/or achieved glycemic control within 24 months of diagnosis. These assessments were conducted nationally and stratified by HIV status.

## Methods

### NHLS cohort creation and description

South Africa’s National Health Laboratory Service (NHLS) is the sole provider of public-sector laboratory services, covering ∼80% of the population across all provinces.^10^ Because patient information linked to laboratory tests is often inconsistently reported or recorded, a single individual may appear under multiple identifiers in the database. To address this, we collaborated with the NHLS to develop a graph-based probabilistic record linkage algorithm that generates a validated, anonymous patient identifier, resulting in the creation of the *NHLS Multi-morbidity Cohort*.^11^ Originally designed to identify unique individuals within the national HIV program, the algorithm was expanded in 2019 to incorporate laboratory tests for HIV, tuberculosis, and non-communicable diseases. In validation, the algorithm achieved a positive predictive value (PPV) of 95.1% and sensitivity of 78.3% when applied to 7,000 non-communicable disease laboratory results from 250 patients—indicating high accuracy in correctly identifying patient matches despite modest sensitivity.^7^

The NHLS Multi-morbidity Cohort now includes >465 million laboratory measurements for ∼50 million unique patients, 22% of whom are <20 years old, with at least one laboratory result recorded between April 1, 2004, and March 31, 2017. For each individual, the dataset captures a unique anonymized identifier, biological sex, age, laboratory test date, test type, result, health facility, district, and province.

### Study population

For this analysis, we included patients from the NHLS Multi-morbidity Cohort if they had an initial blood glucose (either random or fasting) or HbA1c measurement recorded between January 1, 2004, and March 31, 2015. Each of these patients had the opportunity for a follow-up period of 24 months to evaluate T1DM care cascade outcomes. As we did not have a confirmation of T1DM (vs T2DM), we instead used an age proxy and limited our analysis to patients who were <20 years of age when they had their first blood glucose or HbA1c test. Since we lacked data on pregnancy status for female participants, we cannot exclude the possibility that some subjects with gestational diabetes, especially those of reproductive age (15-19 years of age), might be present in our cohort.

### Diabetes guidelines

During the study period, public-sector health facilities in South Africa followed the Society for Endocrinology, Metabolism and Diabetes of South Africa (SEMDSA) 2009^12^ and 2012^13^ diabetes guidelines. T1DM often presents with symptomatic hyperglycemia and, in some cases, diabetic ketoacidosis. Diagnosis is primarily clinical, confirmed by laboratory testing. Typical symptoms include polyuria, polydipsia, unexplained weight loss, polyphagia, visual disturbance, and fatigue, with confirmation using fasting plasma glucose, random plasma glucose, or HbA1c tests.^12,13^ While these tests are recommended for patients presenting with symptoms of diabetes, routine screening of asymptomatic individuals is not common practice.

The SEMDSA guidelines specifically recommend HbA1c and random or fasting plasma glucose tests for diagnosis and monitoring of glycemic control in T1DM. In our cohort, however, random plasma glucose was the predominant diagnostic test used (Supplemental Figure 1a–1f). In practice, clinicians may have relied on point-of-care glucose testing as a substitute for formal laboratory measurements, given the appeal of immediate results. The guidelines emphasize that diagnosis and ongoing monitoring should be based on laboratory-confirmed results rather than point-of-care devices^12,13^ because the latter can introduce measurement error, lack standardization, and are not captured in the NHLS database. This distinction matters both for ensuring clinical accuracy and for interpreting patterns of care in our study, which is limited to laboratory-based results.

### Outcomes

Study outcomes are drawn from the diabetes cascade of care illustrated in Figure 1. We defined three sequential, primary outcomes for the study, as shown below:

**Figure 1.**
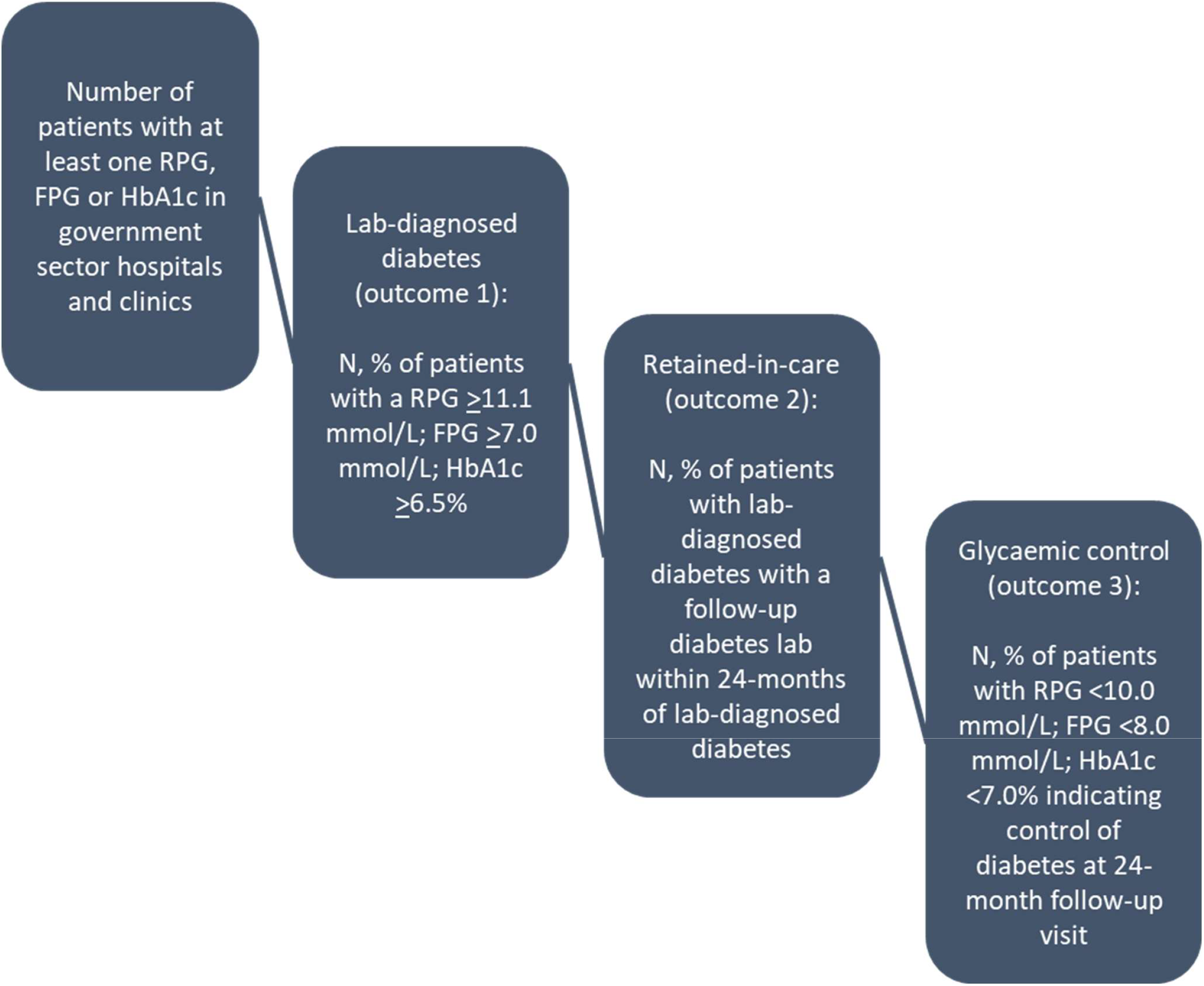
The type 1 diabetes care cascade based on laboratory data.

1. *Laboratory-diagnosed diabetes:* one elevated HbA1c ≥6.5%; or fasting glucose ≥7.0mmol/l; or random glucose ≥11.1mmol/l.^12,13^
2. *Retained-in-care:* an HbA1c, fasting glucose, or random glucose at least one month to up to 24 months after the preliminary laboratory-diagnosis of diabetes. As a sensitivity analysis, we also broadened the definition of retained-in-care to encompass all laboratory tests conducted within 24 months following a diabetes diagnosis— not just those specific to diabetes—to assess if patients were accessing care for other reasons.
3. *Glycaemic control:* an HbA1c <7.0%; or fasting glucose <8.0mmol/l; or random glucose <10.0mmol/l within 24 months of the laboratory-based diabetes diagnosis.^12,13^

The outcomes we assessed align with key steps in the T1DM care continuum, as captured through laboratory data. First, we identified the total number of unique patients who underwent diabetes-related testing. We then estimated the proportion of these patients who: (1) met laboratory diagnostic criteria for T1DM; (2) were retained in care; and (3) achieved glycemic control at their follow-up laboratory test. Importantly, the denominator for each outcome was the population of patients who received diabetes testing, representing only a subset of all prevalent T1DM cases. Results were stratified by facility level (hospital or clinic) and HIV status (defined as patients with an HIV-associated laboratory test – a CD4 count, HIV viral load, or positive HIV test – any time prior to first blood glucose or HbA1c up to 24 months after). As results did not differ by province, we display results at the national level. Estimates of all outcomes were age-standardized to the age distribution of the South African population <20 years of age using mid-year population estimates for 2021^14^ and five-year age-categories.

### Data analysis

We used three mixed effects modified Poisson regression models^15^ to determine predictors of all three outcomes listed above. Facilities nested within districts were included as random effects to control for within-group homogeneity for districts and individual health facilities. In the models we controlled for HIV status, acute tuberculosis infection (defined as patients with a tuberculosis-associated test (i.e. culture, smear, first-line probe assay, drug susceptibility tests, GeneXpert, polymerase chain reaction) 6 months prior to blood glucose or HbA1c and up to 6 months after), province, facility level (hospital or clinic based on location of first blood glucose/HbA1c performed), year of first diabetes test (≤2010, 2011, 2012, 2013, 2014 and 2015), sex, and age (0-4.9, 5-9.9, 10-14.9 and 15-19.9 years). For the model assessing predictors of laboratory-diagnosed T1DM and glycemic control, we also included the type of diagnostic test used, i.e. HbA1c or fasting or plasma glucose.

## Results

### Cohort description and testing trends

Between January 1, 2004, and March 31, 2015, 256,449 individuals underwent testing for diabetes in either a hospital or clinic, as detailed in Table 1. At the time of their first test, the median age of these patients was 4 years (interquartile range (IQR): 0-14 years), and half were female (50.0%). Gauteng Province recorded the youngest median age for patients receiving their first diabetes laboratory test (1 year; IQR: 0-9 years). This is substantially younger than the median ages observed in other provinces, which ranged from 5 to 12 years. Most were living without HIV (86.9%) or acute tuberculosis infection (99.7%). Across all nine provinces, there was consistency in the distributions of gender, median fasting plasma glucose, median random plasma glucose, type of facility, and tuberculosis.

**Table 1.** Demographic and clinical characteristics of the cohort at first blood glucose and HbA_1c_ stratified by province (N=256,449).

| Characteristic | Measure | GP<br>(n= 111,631, 4<br>3.5%) | KZN<br>(n= 14,758,<br>5.8%) | WC<br>(n= 19,584,<br>7.6%) | EC<br>(n= 26,232,<br>10.2%) | FS<br>(n= 31,891,<br>12.4%) | LP<br>(n=19,421,<br>7.6%) | MP<br>(n=9,969, 3.9%) | NC<br>(n=2,501,<br>1.0%) | NW<br>(n=20,462, 8%) | Total<br>(N= 256,449) |
| --- | --- | --- | --- | --- | --- | --- | --- | --- | --- | --- | --- |
| Female | n (%) | 52,407 (46.9%) | 7,968 (54%) | 9,477 (48.4%) | 13,522 (51.5%) | 16,362 (51.3%) | 11,160 (57.5%) | 5,363 (53.8%) | 1,373 (54.9%) | 10,575 (51.7%) | 128,207 (50.0%) |
| Age (years) |  |  |  |  |  |  |  |  |  |  |  |
| 0-4.9 | n (%) | 74,472 (66.7%) | 4,486 (30.4%) | 8,591 (43.9%) | 11,224 (42.8%) | 14,915 (46.8%) | 6,199 (31.9%) | 3,700 (37.1%) | 837 (33.5%) | 10,211 (49.9%) | 134,635 (52.5%) |
| 5-9.9 | n (%) | 9,571 (8.6%) | 2,843 (19.3%) | 2,877 (14.7%) | 3,867 (14.7%) | 3,673 (11.5%) | 2,170 (11.2%) | 1,515 (15.2%) | 337 (13.5%) | 2,456 (12%) | 29,309 (11.4%) |
| 10-14.9 | n (%) | 9,203 (8.2%) | 3,098 (21%) | 3,175 (16.2%) | 3,771 (14.4%) | 4,275 (13.4%) | 2,928 (15.1%) | 1,571 (15.8%) | 441 (17.6%) | 2,435 (11.9%) | 30,897 (12%) |
| 15-19.9 | n (%) | 18,385 (16.5%) | 4,331 (29.3%) | 4,941 (25.2%) | 7,370 (28.1%) | 9,028 (28.3%) | 8,124 (41.8%) | 3,183 (31.9%) | 886 (35.4%) | 5,360 (26.2%) | 61,608 (24%) |
| Age (years) | median (IQR) | 1 (0, 9) | 10 (3, 15) | 7 (1, 15) | 7 (1, 15) | 6 (0, 15) | 12 (2, 17) | 9 (2, 16) | 10 (2, 17) | 5 (0, 15) | 4 (0, 14) |
| lab-diagnosed diabetes |  | 12,612 (11.3%) | 2,139 (14.5%) | 2,139 (10.9%) | 3,212 (12.2%) | 2,039 (6.4%) | 2,812 (14.5%) | 1,689 (16.9%) | 309 (12.4%) | 2,128 (10.4%) | 29,079 (11.3%) |
| lab-diagnosed pre-diabetes |  | 9,913 (8.9%) | 996 (6.7%) | 1,604 (8.2%) | 2,047 (7.8%) | 2,190 (6.9%) | 1,605 (8.3%) | 824 (8.3%) | 167 (6.7%) | 1,443 (7.1%) | 20,789 (8.1%) |
| FPG (mmol/l) | median (IQR) | 4.8 (4.2, 5.9) | 4.7 (4.2, 5.4) | 4.8 (4.3, 5.3) | 4.9 (4.3, 5.9) | 4.7 (4.1, 5.5) | 5.0 (4.2, 6.9) | 4.9 (4.3, 6.1) | 4.6 (4.2, 5.3) | 4.8 (4.2, 5.7) | 4.8 (4.2, 5.7) |
| RPG (mmol/l) | median (IQR) | 5.3 (4.4, 7.0) | 4.8 (4.3, 5.6) | 5.5 (4.6, 7.2) | 5.3 (4.5, 6.9) | 5.0 (4.1, 6.2) | 5.4 (4.4, 7.3) | 5.3 (4.5, 7.3) | 5.2 (4.4, 6.7) | 5.1 (4.2, 6.6) | 5.3 (4.4, 6.8) |
| HbA<sub>1c</sub> (%) | median (IQR) | 6.0 (5.4, 9.7) | 6.8 (5.6, 11.6) | NA | 5.9 (5.3, 11.0) | 5.6 (5.1, 9.3) | 9.8 (5.9, 14.3) | 7.0 (5.6, 12.0) | 5.8 (5.2, 8.5) | 6.4 (5.5, 11.4) | 6.3 (5.5, 11.2) |
| HIV status |  |  |  |  |  |  |  |  |  |  |  |
| HIV- |  | 99,738 (89.3%) | 8,219 (55.7%) | 17,737 (90.6%) | 22,506 (85.8%) | 28,230 (88.5%) | 18,379 (94.6%) | 7,728 (77.5%) | 2,293 (91.7%) | 17,942 (87.7%) | 222,772 (86.9%) |
| HIV+ |  | 11,893 (10.7%) | 6,539 (44.3%) | 1,847 (9.4%) | 3,726 (14.2%) | 3,661 (11.5%) | 1,042 (5.4%) | 2,241 (22.5%) | 208 (8.3%) | 2,520 (12.3%) | 33,677 (13.1%) |
| Tuberculosis |  |  |  |  |  |  |  |  |  |  |  |
| positive |  | 111,346 (99.7%) | 14,614 (99.0%) | 19,458 (99.4%) | 26,173 (99.8%) | 31,803 (99.7%) | 19,407 (99.9%) | 9,948 (99.8%) | 2,493 (99.7%) | 20,441 (99.9%) | 255,683 (99.7%) |
| negative |  | 285 (0.3%) | 144 (1.0%) | 126 (0.6%) | 59 (0.2%) | 88 (0.3%) | 14 (0.1%) | 21 (0.2%) | 8 (0.3%) | 21 (0.1%) | 766 (0.3%) |
| Facility type |  |  |  |  |  |  |  |  |  |  |  |
| hospital |  | 109,481 (98.1%) | 13,663 (92.6%) | 18,116 (92.5%) | 24,014 (91.5%) | 30,307 (95.0%) | 17,127 (88.2%) | 9,062 (90.9%) | 1,923 (76.9%) | 18,874 (92.2%) | 242,567 (94.6%) |
| clinic |  | 2,150 (1.9%) | 1,095 (7.4%) | 1,468 (7.5%) | 2,218 (8.5%) | 1,584 (5.0%) | 2,294 (11.8%) | 907 (9.1%) | 578 (23.1%) | 1,588 (7.8%) | 13,882 (5.4%) |
| Year |  |  |  |  |  |  |  |  |  |  |  |
| 2004 |  | 8,118 (7.3%) | - | 3,330 (17%) | 2,789 (10.6%) | 3,040 (9.5%) | 2,090 (10.8%) | 993 (10%) | 268 (10.7%) | 2,400 (11.7%) | 23,028 (9%) |
| 2005 |  | 11,353 (10.2%) | - | 2,608 (13.3%) | 3,851 (14.7%) | 3,289 (10.3%) | 2,737 (14.1%) | 1,442 (14.5%) | 148 (5.9%) | 3,754 (18.3%) | 29,182 (11.4%) |
| 2006 |  | 12,995 (11.6%) | - | 2,312 (11.8%) | 3,852 (14.7%) | 3,640 (11.4%) | 2,786 (14.3%) | 1,106 (11.1%) | 262 (10.5%) | 3,548 (17.3%) | 30,501 (11.9%) |
| 2007 |  | 12,397 (11.1%) | - | 1,854 (9.5%) | 2,767 (10.5%) | 3,778 (11.8%) | 2,519 (13%) | 1,108 (11.1%) | 195 (7.8%) | 2,291 (11.2%) | 26,909 (10.5%) |
| 2008 |  | 12,720 (11.4%) | - | 1,225 (6.3%) | 2,731 (10.4%) | 3,833 (12%) | 2,273 (11.7%) | 1,116 (11.2%) | 260 (10.4%) | 2,692 (13.2%) | 26,850 (10.5%) |
| 2009 |  | 12,160 (10.9%) | - | 1,466 (7.5%) | 2,256 (8.6%) | 3,789 (11.9%) | 2,051 (10.6%) | 1,036 (10.4%) | 294 (11.8%) | 1,756 (8.6%) | 24,808 (9.7%) |
| 2010 |  | 12,043 (10.8%) | 2,594 (17.6%) | 1,392 (7.1%) | 2,100 (8%) | 3,594 (11.3%) | 1,752 (9%) | 1,016 (10.2%) | 276 (11%) | 1,421 (6.9%) | 26,188 (10.2%) |
| 2011 |  | 10,658 (9.5%) | 3,001 (20.3%) | 1,353 (6.9%) | 1,765 (6.7%) | 2,860 (9%) | 1,312 (6.8%) | 826 (8.3%) | 250 (10%) | 1,340 (6.5%) | 23,365 (9.1%) |
| 2012 |  | 7,363 (6.6%) | 2,714 (18.4%) | 1,258 (6.4%) | 1,449 (5.5%) | 1,900 (6%) | 471 (2.4%) | 325 (3.3%) | 160 (6.4%) | 321 (1.6%) | 15,961 (6.2%) |
| 2013 |  | 4,439 (4%) | 2,205 (14.9%) | 1,234 (6.3%) | 1,187 (4.5%) | 1,085 (3.4%) | 438 (2.3%) | 389 (3.9%) | 119 (4.8%) | 312 (1.5%) | 11,408 (4.4%) |
| 2014 |  | 5,454 (4.9%) | 2,887 (19.6%) | 1,243 (6.3%) | 1,147 (4.4%) | 869 (2.7%) | 670 (3.4%) | 470 (4.7%) | 175 (7%) | 457 (2.2%) | 13,372 (5.2%) |
| 2015 |  | 1,931 (1.7%) | 1,357 (9.2%) | 309 (1.6%) | 338 (1.3%) | 214 (0.7%) | 322 (1.7%) | 142 (1.4%) | 94 (3.8%) | 170 (0.8%) | 4,877 (1.9%) |

\*FPG - fasting blood glucose; RPG - random blood glucose; HbA<sub>1c</sub> - glycated hemoglobin A<sub>1c</sub>; GP – Gauteng; KZN – KwaZulu Natal; WC – Western Cape; EC – Eastern Cape; FS – Free State; LP – Limpopo; MP – Mpumalanga; NC – Northern Cape; NW – North West

The median HbA1c value for the entire sample was 6.3% (IQR: 5.5, 11.2%). In the provincial breakdown, Limpopo Province reported the highest median HbA1c value, which was 9.8% (IQR: 5.9-14.3%). In comparison, median values for the remaining provinces varied between 5.6% and 7.0%. The Western Cape Province did not utilize HbA1c tests in clinics during the specified time period. HIV prevalence for the cohort was 13.1%. KwaZulu Natal and Mpumalanga were highest at 44.3% and 22.5% of youth living with HIV (YLWH), respectively, while Limpopo (5.4%) and Northern Cape (8.3%) had the lowest HIV prevalence.

Supplemental Figures 1a-1f provide a visual representation of blood glucose and HbA1c testing occurrences over quarterly intervals. These figures differentiate results by the level of healthcare facility and HIV status. Notably, each patient could be represented multiple times if they underwent more than one test. The predominant type of test used was random plasma glucose, which accounted for 84.3% (467,746/554,817) of tests. From 2010 onwards, there was a noticeable increase in the use of HbA1c tests. The majority of diabetes tests (94.7%; 525,595/554,817 tests), were conducted in a hospital setting (Supplemental Figures 1a-1c), while the volume of patients undergoing diabetes testing in primary healthcare clinics was considerably lower, accounting for only 5.3% (29,222/554,817 tests).

### T1DM care cascade

Figure 2 and Table 2 provide an overall comprehensive view of the age-standardized laboratory-based T1DM care cascade in South Africa between 2012 and 2017. Of the total 256,449 patients screened for diabetes during this period, 31,010 (12.1%; 95% CI: 12.0-12.2%) exhibited HbA1c or blood glucose laboratory values indicative of T1DM. Among those with a laboratory diagnosis, 4,934 individuals (15.9%; 95% CI: 15.5-16.3%) were retained in care 24 months after having a lab-diagnosis of diabetes. A smaller proportion of those with a laboratory-based diagnosis (n=2,239; 7.2%; 95% CI: 6.9-7.5%) managed to achieve glycaemic control within the 24-months of their diagnosis of T1DM. In the conditional cascade (in which each step is conditional on achieving the previous step), of these individuals retained in care, 45.4% (95% CI: 44.0-46.8%) successfully achieved glycemic control within 24 months.

**Table 2.** The type 1 diabetes care cascade overall and the transitions between stages stratified by facility type (hospital and clinic) and HIV status (N=256,449).

|  | No. patients<br>tested<br>n (%; 95% CI) | Lab-diagnosed<br>diabetes<br>n (%; 95% CI) | Retained-in-care<br>within 24-mo.<br>n (%; 95% CI) | Controlled diabetes<br>within 24-mo.<br>n (%; 95% CI) |
| --- | --- | --- | --- | --- |
| **National (n=256,449)** |  |  |  |  |
| YLWH overall | 33,677 | 1,989 (5.9%; 5.7-6.2%) | 335 (16.8%; 15.3-18.6%) | 243 (12.2%; 10.9-13.7%) |
| YLWOH overall | 222,772 | 29,021 (13%; 12.9-13.2%) | 4,599 (15.8%; 15.4-16.3%) | 1,996 (6.9%; 6.6-7.2%) |
| Total overall | 256,449 | 31,010 (12.1%; 12-12.2%) | 4,934 (15.9%; 15.5-16.3%) | 2,239 (7.2%; 6.9-7.5%) |
| YLWH transition between stages | 33,677 | 1,989 (5.9%; 5.7-6.2%) | 335 (16.8%; 15.3-18.6%) | 243 (72.5%; 67.5-77%) |
| YLWOH transition between stages | 222,772 | 29,021 (13%; 12.9-13.2%) | 4,599 (15.8%; 15.4-16.3%) | 1,996 (43.4%; 42-44.8%) |
| Total transition between stages | 256,449 | 31,010 (12.1%; 12-12.2%) | 4,934 (15.9%; 15.5-16.3%) | 2,239 (45.4%; 44-46.8%) |
| **Hospital (n=242,567)** |  |  |  |  |
| YLWH overall | 30,631 | 1,947 (6.4%; 6.1-6.6%) | 323 (16.6%; 15-18.3%) | 239 (12.3%; 10.9-13.8%) |
| YLWOH overall | 211,936 | 27,642 (13%; 12.9-13.2%) | 4,442 (16.1%; 15.6-16.5%) | 1,948 (7%; 6.8-7.4%) |
| Total overall | 242,567 | 29,589 (12.2%; 12.1-12.3%) | 4,765 (16.1%; 15.7-16.5%) | 2187 (7.4%; 7.1-7.7%) |
| YLWH transition between stages | 30,631 | 1,947 (6.4%; 6.1-6.6%) | 323 (16.6%; 15-18.3%) | 239 (74%; 68.9-78.5%) |
| YLWOH transition between stages | 211,936 | 27,642 (13%; 12.9-13.2%) | 4,442 (16.1%; 15.6-16.5%) | 1,948 (43.9%; 42.4-45.3%) |
| Total transition between stages | 242,567 | 29,589 (12.2%; 12.1-12.3%) | 4,765 (16.1%; 15.7-16.5%) | 2,187 (45.9%; 44.5-47.3%) |
| **Clinic (n=13,882)** |  |  |  |  |
| YLWH overall | 3,046 | 42 (1.4%; 1-1.9%) | 12 (28.6%; 17.2-43.6%) | 4 (9.5%; 3.8-22.1%) |
| YLWOH overall | 10,836 | 1,379 (12.7%; 12.1-13.4%) | 157 (11.4%; 9.8-13.2%) | 48 (3.5%; 2.6-4.6%) |
| Total overall | 13,882 | 1,421 (10.2%; 9.7-10.8%) | 169 (11.9%; 10.3-13.7%) | 52 (3.7%; 2.8-4.8%) |
| YLWH transition between stages | 3,046 | 42 (1.4%; 1-1.9%) | 12 (28.6%; 17.2-43.6%) | 4 (33.3%; 13.8-60.9%) |
| YLWOH transition between stages | 10,836 | 1,379 (12.7%; 12.1-13.4%) | 157 (11.4%; 9.8-13.2%) | 48 (30.6%; 23.9-38.2%) |
| Total transition between stages | 13,882 | 1,421 (10.2%; 9.7-10.8%) | 169 (11.9%; 10.3-13.7%) | 52 (30.8%; 24.3-38.1%) |

\*YLWH – youth living with HIV; YLWOH – youth living without HIV

**Figure 2.**
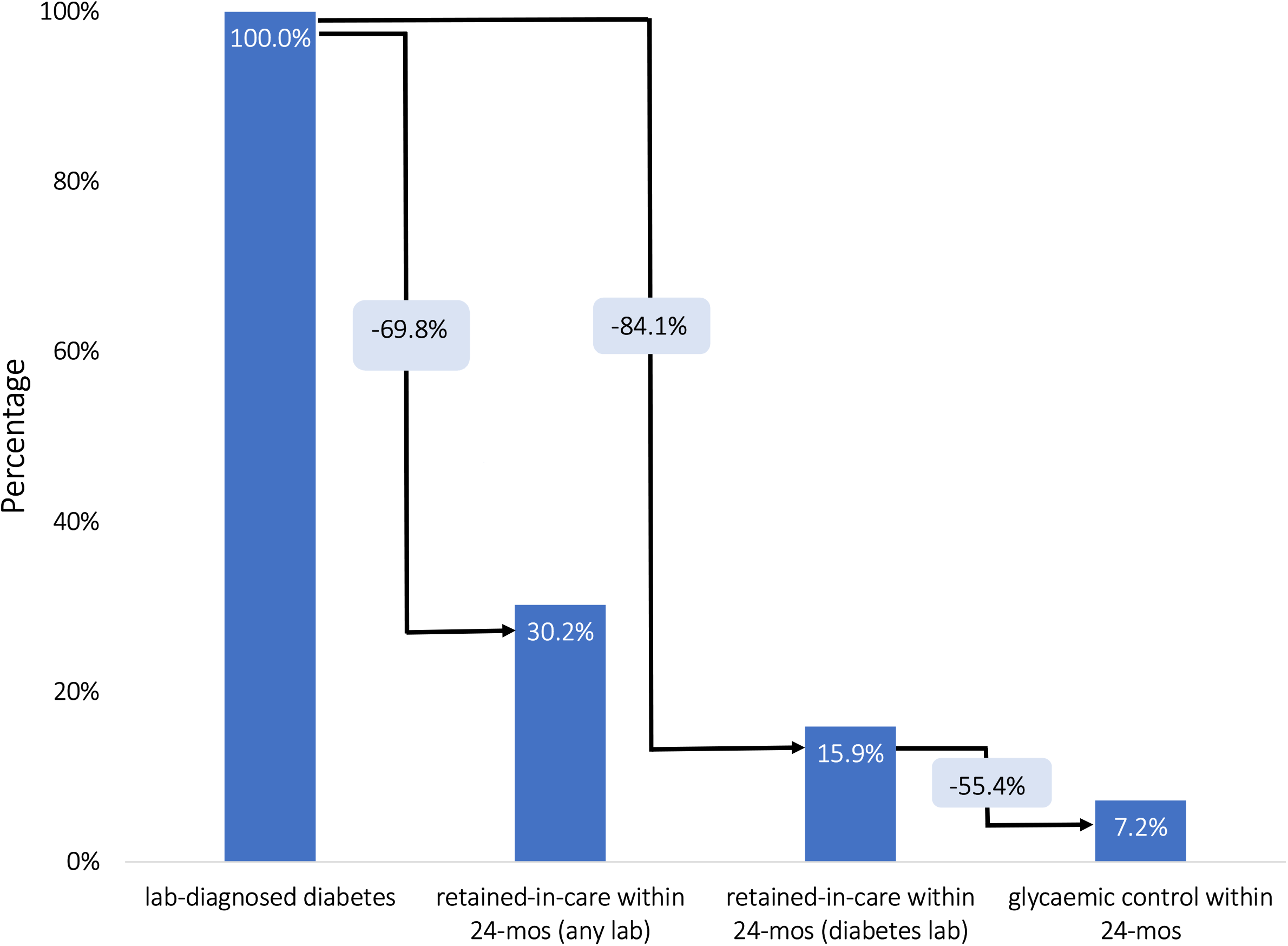
The type 1 diabetes care cascade based on laboratory data (N=275,081).

When we included all types of laboratory tests conducted within 24 months after diagnosis—rather than restricting to diabetes-specific tests—a larger proportion of patients with a laboratory diagnosis of T1DM appeared to be seeking medical care for other health-related issues. Of the 31,010 patients diagnosed with diabetes, 9,351 (30.2%; 29.6–30.7%) underwent any form of laboratory test in the ensuing 24 months. In contrast, only 4,934 (15.9%; 95% CI: 15.5–16.3%) received a diabetes-specific laboratory test during the same period, leaving a gap of 4,417 patients. The majority of these patients were tested for hemoglobin, alanine transaminase, and HIV. This pattern matters because it suggests that many individuals with T1DM continued to access health services but not for diabetes-related monitoring—either because co-morbid conditions (e.g., HIV) prompted ongoing testing, or because diabetes care was de-prioritized once patients were engaged in the health system. As such, these findings highlight potential missed opportunities to integrate diabetes management into routine care encounters.

### Hospital vs. clinic

We found that those being tested for diabetes in a hospital setting had a lower median age (2 years (IQR: 0-13 years) compared to those tested in clinics (12 years IQR: 1-17 years) (Supplemental Table 1). Table 2 provides a breakdown of transitions between stages of the diabetes care cascade by facility level. In clinics, 13,882 individuals were tested for diabetes, with 10.2% (95% CI: 9.7-10.8%) being diagnosed with T1DM. This rate is similar to hospitals, which diagnosed 12.2% (95% CI: 12.1-12.3%) of the 242,567 individuals tested. Post-diagnosis, 11.9% (95% CI: 10.3-13.7%) of individuals in clinics were retained in care, which is a slightly lower than the 16.1% (95% CI: 15.7-16.5%) in hospitals. Only 3.7% (95% CI: 2.8-4.8%) of those in clinics achieved glycemic control within 24 months, while hospitals reported a slightly higher rate of 7.4% (95% CI: 7.1-7.7%). When evaluating the transition from retention in care to achieving glycemic control, hospitals also fared better than clinics, with 45.9% (95% CI: 44.5-47.3%) meeting glycemic targets versus 30.8% (95% CI: 24.3-38.1%), respectively.

### Youth living with HIV (YLWH) vs youth living without HIV (YLWOH)

We found that YLWH had a lower median age (1 year (IQR: 0-9 years) at laboratory diagnosis of T1DM than did YLWOH (2 years IQR: 0-13 years) (Supplemental Table 1). In Table 2, among the 222,772 YLWOH tested for diabetes, 13.0% (95% CI: 12.9-13.2%) were diagnosed with the condition. In comparison, of the 33,677 YLWH tested, only 5.9% (95% CI: 5.7-6.2%) were diagnosed. Both groups displayed a similar proportion retained in care post-diagnosis. YLWH had a higher proportion achieving glycemic control (12.2%; 95% CI: 10.9-13.7%) than did YLWOH (6.9%; 95% CI: 6.6-7.2%). When assessing the progression from retention in care to glycemic control, 72.5% (95% CI: 67.5-77.0%) of YLWH and 43.4% (95% CI: 42.0-44.8%) of YLWOH achieved this outcome.

### Predictors of laboratory-diagnosed T1DM

Table 4 presents findings from the mixed effects Poisson regression analysis. As age increased, the likelihood of receiving a laboratory diagnosis for T1DM decreased. Females had a slightly higher likelihood of being diagnosed than males. YLWH had a 53% reduced risk (adjusted Risk Ratio (aRR): 0.53; 95% CI: 0.50-0.57) of receiving a laboratory diagnosis for T1DM than did YLWOH. Those with an acute tuberculosis infection had a 12% (but imprecise) lowered risk (aRR: 0.88; 95% CI: 0.62-1.23) of a T1DM diagnosis compared to individuals without tuberculosis. When considering the facility at which the diagnosis occurred, those tested at clinics had a 23% reduced risk (aRR: 0.77; 95% CI: 0.70-0.84) of being diagnosed with diabetes than did those tested at hospitals. The data also showed temporal trends in the risk of diabetes diagnosis. Compared to the years before 2010, risk decreased by 12% (aRR: 0.88; 95% CI: 0.84-0.93) in 2011, 26% (aRR: 0.74; 95% CI: 0.70, 0.79) in 2012, 29% (aRR: 0.71; 95% CI: 0.66, 0.76) in 2013, 28% (aRR: 0.72; 95% CI: 0.67-0.77) in 2014, and 30% (aRR: 0.7; 95% CI: 0.64-0.78) in 2015. Lastly, relative to those tested with HbA1c, individuals diagnosed via fasting plasma glucose and random plasma glucose tests had a significantly reduced likelihood of receiving a laboratory-confirmed diagnosis of T1DM (aRR: 0.12; 95% CI: 0.11–0.13 and aRR: 0.24; 95% CI: 0.22–0.26, respectively).

**Table 4.** Crude and adjusted nested models for the outcomes of laboratory-diagnosed diabetes, remained in care and controlled diabetes in the national cohort (N=256,449).

| Characteristic | Measure | Lab-diagnosed diabetes |  | Remained in care |  | Glycaemic control |  |
| --- | --- | --- | --- | --- | --- | --- | --- |
|  |  | crude RR<br>(95% CI) | adjusted RR<br>(95% CI) | crude RR<br>(95% CI) | adjusted RR<br>(95% CI) | crude RR<br>(95% CI) | adjusted RR<br>(95% CI) |
| Gender | Male | ref. | ref. | ref. | ref. | ref. | ref. |
|  | Female | 1.08 (1.05, 1.11) | 1.09 (1.06, 1.12) | 1.05 (0.97, 1.14) | 0.98 (0.90, 1.06) | 0.91 (0.81, 1.02) | 0.96 (0.85, 1.08) |
| Test type | HbA1c | ref. | ref. | - | - | ref. | ref. |
|  | RPG | 0.16 (0.15, 0.17) | 0.12 (0.11, 0.13) | - | - | 3.51 (2.71, 4.55) | 3.10 (2.11, 4.55) |
|  | FPG | 0.30 (0.28, 0.32) | 0.24 (0.22, 0.26) | - | - | 3.26 (2.01, 5.28) | 3.03 (1.76, 5.23) |
| Age (years) | 0-4.9 | ref. | ref. | ref. | ref. | ref. | ref. |
|  | 5-9.9 | 0.60 (0.57, 0.63) | 0.58 (0.55, 0.61) | 1.57 (1.34, 1.83) | 1.57 (1.35, 1.84) | 0.84 (0.68, 1.02) | 0.93 (0.76, 1.14) |
|  | 10-14.9 | 0.76 (0.73, 0.8) | 0.67 (0.64, 0.70) | 3.18 (2.85, 3.56) | 3.23 (2.89, 3.62) | 0.55 (0.47, 0.66) | 0.61 (0.52, 0.72) |
|  | 15-19.9 | 0.62 (0.60, 0.64) | 0.55 (0.53, 0.57) | 2.59 (2.32, 2.88) | 2.58 (2.32, 2.88) | 0.53 (0.44, 0.62) | 0.58 (0.49, 0.69) |
| HIV status | YLWOH | ref. | ref. | ref. | ref. | ref. | ref. |
|  | YLWH | 0.46 (0.44, 0.49) | 0.53 (0.50, 0.57) | 1.77 (1.55, 2.03) | 1.86 (1.62, 2.13) | 1.46 (1.24, 1.72) | 1.25 (1.06, 1.48) |
| Tuberculosis status | Tuberculosis - | ref. | ref. | ref. | ref. | ref. | ref. |
|  | Tuberculosis + | 0.63 (0.45, 0.88) | 0.88 (0.62, 1.23) | 2.14 (1.21, 3.8) | 1.74 (0.97, 3.10) | 0.87 (0.28, 2.72) | 0.73 (0.23, 2.29) |
| Facility type at first test | Hospital | ref. | ref. | ref. | ref. | ref. | ref. |
|  | Clinic | 1.02 (0.91, 1.14) | 0.77 (0.70, 0.84) | 0.64 (0.50, 0.82) | 0.57 (0.44, 0.73) | 0.65 (0.39, 1.08) | 1.02 (0.61, 1.73) |
| Year | ≤2010 | ref. | ref. | ref. | ref. | ref. | ref. |
|  | 2011 | 0.94 (0.89, 0.98) | 0.88 (0.84, 0.93) | 0.81 (0.69, 0.96) | 0.81 (0.69, 0.96) | 0.85 (0.66, 1.10) | 0.94 (0.73, 1.20) |
|  | 2012 | 1.05 (0.99, 1.11) | 0.74 (0.70, 0.79) | 1.04 (0.87, 1.24) | 1.00 (0.84, 1.19) | 0.81 (0.62, 1.05) | 1.1 (0.84, 1.43) |
|  | 2013 | 1.38 (1.30, 1.47) | 0.71 (0.66, 0.76) | 1.41 (1.21, 1.66) | 1.36 (1.16, 1.59) | 0.51 (0.37, 0.70) | 0.97 (0.67, 1.39) |
|  | 2014 | 1.28 (1.21, 1.36) | 0.72 (0.67, 0.77) | 1.10 (0.93, 1.29) | 1.04 (0.88, 1.23) | 0.44 (0.32, 0.62) | 0.81 (0.56, 1.18) |
|  | 2015 | 1.09 (0.99, 1.20) | 0.70 (0.64, 0.78) | 1.18 (0.91, 1.53) | 1.06 (0.82, 1.39) | 0.36 (0.19, 0.67) | 0.80 (0.40, 1.60) |

FPG - fasting plasma glucose; RPG - random plasma glucose; HbA1c - glycated hemoglobin A1c; all models are adjusted for province.

### Predictors of retention in care for T1DM

Table 4 also indicates that among those diagnosed with T1DM, older children and adolescents were more likely to continue to be retained in care than individuals <5 years of age. Notably, YLWH had a higher propensity to remain in care (aRR: 1.86; 95% CI: 1.62-2.13) compared to YLWOH. Patients who were diagnosed in clinics showed a significant 43% reduced risk (aRR: 0.57; 95% CI: 0.44-0.73) of being retained in care than did those diagnosed in hospitals. The data also highlighted a temporal trend in retention in care. There was an upward trend in the likelihood of individuals being retained in care in the years after 2010. We saw little difference in this outcome by gender.

### Predictors of glycaemic control

Finally, Table 4 also outlines determinants of glycemic control among patients who continued with their care. Patients ≥5 had a reduced likelihood, ranging from 7% to 42%, of achieving glycemic control in comparison to those <5 years old. As with retention, YLWH were 25% more likely (aRR: 1.25; 95% CI: 1.06-1.48) to attain glycemic control than YLWOH. A noteworthy trend is the consistent decline in glycemic control rates after 2010 compared to in earlier years. Those patients diagnosed with diabetes using fasting plasma glucose and random plasma glucose tests had a higher likelihood (by 203% and 210%, respectively) of reaching glycemic control than those identified through the HbA1c test. We saw little difference in this outcome by gender and healthcare facility level.

## Discussion

To our knowledge, our study is the first of its kind in sub-Saharan Africa to prospectively evaluate the T1DM care continuum at a national level. We uncovered substantial shortcomings in the retention of patients and in attaining glycemic control within a 24-month span following a laboratory diagnosis of T1DM. These findings are consistent with our earlier research on the T2DM care cascade, also based on NHLS data^7^, and support existing literature that indicates diabetes management in sub-Saharan African countries is inadequate.^5,6^

In 2021, the International Diabetes Federation reported a total of 59,500 (4,526 (7.6%) in South Africa), total cases of T1DM in children and adolescents globally, representing a regional prevalence of <1%, consistent with previous reports from East Africa.^1,16-18^ These numbers likely underrepresent the actual prevalence of T1DM in the region due to the lack of comprehensive data and the high mortality rates linked to the condition. A longitudinal study over two decades at Chris Baragwanath Academic Hospital in Soweto, South Africa, which included 88 patients with T1DM, showed a stark 43% crude mortality rate.^19^ It is reported that within sub-Saharan Africa many children with T1DM may die before they are diagnosed^19^ so it is possible that our data under-estimate the true number of cases

Due to the absence of prior research on retention rates in diabetes care for children and adolescents, it may be useful to draw a parallel with retention in HIV care—a reasonable analogue because both conditions necessitate lifelong treatment are likely to be fatal without treatment. According to a meta-analysis of cohorts from sub-Saharan Africa, the retention rate in pediatric HIV care was 77% by 36 months.^20^ We found that only 15.9% of children and adolescents with a laboratory-confirmed diagnosis of T1DM were retained in care over a 24 month follow-up period, substantially lower than retention in HIV care. This retention rate was also lower than the 30.6% estimated from NHLS data when assessing T2DM care among individuals greater than 30 years of age.^7^ Expanding our definition of ‘retention in care’ to include any laboratory test, not solely diabetes-specific ones, we found that 30.2% of diagnosed patients continued to engage with healthcare services. Within this group, 47.2% had other medical tests conducted but did not receive any further diabetes-specific follow-up. This finding underscores the critical gaps in the healthcare continuum for T1DM patients and suggests a need for more targeted interventions to improve retention in care.

For individuals with T1DM, it is crucial to maintain glycemic control via consistent insulin therapy. A 2020 study across 12 African countries reported that only 16.6% of 712 young adults with T1DM achieved target HbA1c levels, with one fourth hospitalized in the past year due to diabetic ketoacidosis (58.1%) or hypoglycemia (31.1%) (20). In Tanzania, 42.2% of children and young adults with T1DM had recorded HbA1c levels, of which 36% showed levels above 12.5%, far exceeding the optimal threshold of 7.0%. These individuals also had a high incidence of complications including retinopathy (21.5%) and neuropathy (29.4%).^1,16-18^ Our study reveals an even lower rate of glycemic control, with just 7.2% of individuals with T1DM achieving it over 24 months, comparable to what we found in our T2DM care cascade estimates.^7^ This contrasts with higher rates from cross-sectional studies^1,16-18,21^, suggesting longitudinal analysis might provide a more accurate picture of long-term management success.

Access to insulin in sub-Saharan Africa is hindered by the absence of structured management programs and multifaceted healthcare delivery challenges.^22^ These include limited awareness of T1DM, delayed diagnosis, insufficient diagnostic resources, and logistical hurdles in sample transport and processing.^22^ South African public healthcare, the main provider for diabetes care, struggles with overcrowding, leading to excessive wait times for routine treatments. The high cost of managing T1DM, particularly the cost of insulin, imposes substantial financial stress on both patients and the healthcare system.^22^ The convergence of these difficulties and high rates of patient drop-out underscores the urgent need for reforms aimed at improving patient retention and streamlining continuous, effective care.

YLWH are at risk for various HIV-related comorbidities, including cardiovascular diseases, diabetes, liver diseases, renal diseases, and bone disorders, due to the long-term immunological effects of HIV and antiretroviral therapy (ART).^23^ Our analysis shows that YLWH had a 5.9% likelihood of testing positive for diabetes, substantially lower than the 13.0% observed in YLWOH. These results are similar to what we observed in the T2DM analysis using the NHLS data^7^ and could reflect higher levels of mortality in those with both diseases. Research on the relationship between HIV and diabetes in sub-Saharan Africa, particularly among children and young adults, is sparse and has yielded varied results^24^, and it requires further investigation.

We found that YLWH were identified with T1DM at an earlier age in comparison to their counterparts without HIV. This observation suggests several interconnected factors influencing the earlier diagnosis in YLWH. Regular and comprehensive medical examinations, a standard aspect of HIV care, could contribute to an increased rate of early diabetes detection, mirroring findings in adult HIV populations.^24,25^ Additionally, the complex interaction between HIV-induced immune dysfunction, which often involves chronic inflammation and possible autoimmune alterations, may precipitate the earlier development of T1DM.^26,27^ The role of antiretroviral treatments, some of which are linked to metabolic changes, may further influence the risk and timing of a diabetes diagnosis.^28,29^ These considerations highlight the critical need for thorough diabetes screening protocols in YLWH, which could lead to more proactive diabetes management and enrich our understanding of the disease’s pathophysiology in this population.

Patients grappling with multiple health issues, particularly those necessitating diverse treatment strategies, face significant healthcare challenges. This situation is particularly acute among YLWH, who are at an elevated risk for other health problems but frequently lack proper diagnosis and management for them. A study in South Africa found that while 55% of YLWH suffered from at least one additional health issue, very few underwent treatment for these conditions.^23^ Routine healthcare visits offer invaluable opportunities for disease screening and management. YLWH routinely visiting healthcare providers for their antiretroviral treatment could also be assessed and treated for diabetes during these visits. It has been suggested that YLWH undergoing antiretroviral treatment may manage diabetes more effectively than the broader population, but there is currently little evidence to support this. We found that YLWH were somewhat more successful at diabetes management 24 months after their initial diagnosis compared to their non-HIV-infected counterparts. Nevertheless, the overall successful management rates for diabetes were strikingly low.

Our study’s key advantage lies in the extensive scope of our national cohort, encompassing 256,449 individuals. Nevertheless, there are significant limitations to our data. First, the probabilistic matching approach used to compile our cohort could lead to over-matching or under-matching, potentially affecting the accuracy of our findings. We note that our results excluded patients linked to laboratory results deemed unreliable, however, which increases the likelihood that our main findings are robust. For example, considering potential under-matching, our sensitivity analysis indicates that the most by which we could be underestimating retention in care is 22% (1.0-0.78). Adjusting our figures by 28% (1/0.78) would still not substantially alter the overall conclusions of our study. Second, since our evaluation of the care cascade begins with individuals already screened for diabetes, we cannot account for attrition that may occur at the screening phase. This might lead us to overestimate the cascade’s efficacy compared to studies that include screening-stage losses. We chose not to calculate screening rates within this study, as any attempt to do so would be highly speculative. Third, as mentioned above, the denominator for our analysis was limited to those who underwent testing for diabetes due to clinical suspicion or family history. Our results should not be interpreted as population prevalences of T1DM, but rather as estimates derived from individuals who accessed laboratory testing within the public sector. Finally, our analysis may be limited by uncontrolled confounding since the laboratory data does not include detailed clinical factors at the patient level. A linkage of laboratory data with patient-level clinical records would enhance our analysis by providing additional data for screening and further confounding factors.

## Conclusion

Using South Africa’s National Health Laboratory Service database, we found stark gaps in T1DM care: 80% of diagnosed individuals under the age of 20 fell out of regular care, and fewer than 10% maintained glycemic control after two years. Disparities in diabetes prevalence between YLWH and YLWOH require further investigation. Our findings advocate for urgent, refined approaches to T1DM care in South Africa, emphasizing the need to tailor healthcare to improve outcomes.

## Data Availability

The data in our study are not publicly available due to the terms of our contract with the NHLS, but are available from the NHLS with reasonable request.

https://www.nhls.ac.za/

## Declarations

### Ethics approval and consent for participation

Approval for analysis of de-identified NHLS data was granted by Boston University’s Institutional Review Board (protocol no. H-41152), Human Research Ethics Committee of the University of the Witwatersrand (protocol no. M200851) and NHLS Academic Affairs and Research Management System (protocol no. PR2232386). We confirm that all methods were carried out in accordance with relevant guidelines and regulations. Our research was observational, not experimental, in nature. We conducting our analysis using existing NHLS laboratory data from South Africa that is used for routine clinical care of patients in government sector health facilities. We had no contact with patients and no experiments were conducted on humans or human tissues, as such, informed consent was not obtained from subjects and/or their legal guardian(s). An inform consent waiver was received from the Boston University’s Institutional Review Board and the Human Research Ethics Committee of the University of the Witwatersrand.

### Consent for publication

Not applicable

### Competing interests

Authors declare no competing interests.

### Funding

National Institute of Diabetes and Digestive and Kidney Diseases K01DK116929

### Authors’ contributions

All authors were involved in the design of the study. ATB and EL accessed and verified the data. EL did the statistical analysis with support from ATB. EL and ATB wrote the first draft of the article. ATB, EL, NJC, JAG, JB, SK, and MPF have full access to all the data in the study. ATB, JB, JAG, KC, KM, AK, SR, FR, PH, MPF and NJC critically interpreted the results and developed the report. All authors reviewed and approved the final version.

## Acknowledgments

The NIDDK had no role in study design, data collection and analysis, decision to publish, or preparation of the manuscript.

## Authors’ information

Alana T Brennan.

Evelyn Lauren.

Jacob Bor.

Jaya A George

Kamy Chetty.

Koleka Mlisana.

Siyabonga Khoza.

Sydney Rosen.

Frederick Raal.

Patricia Hibberd.

Matthew P Fox.

Nigel J Crowther.

## Notes

### Competing Interest Statement

The authors have declared no competing interest.

### Summary of Updates

No revisions have been made.

