## Supplemental Figure 1 for "Gaps in the Type 1 Diabetes Mellitus care cascade: a national perspective using South Africa’s National Health Laboratory Service (NHLS) database"

**Supplemental Figure 1a-1f. Quarterly glucose (random or fasting) and HbA1c lab events nationally and stratified by HIV status and facility type**.

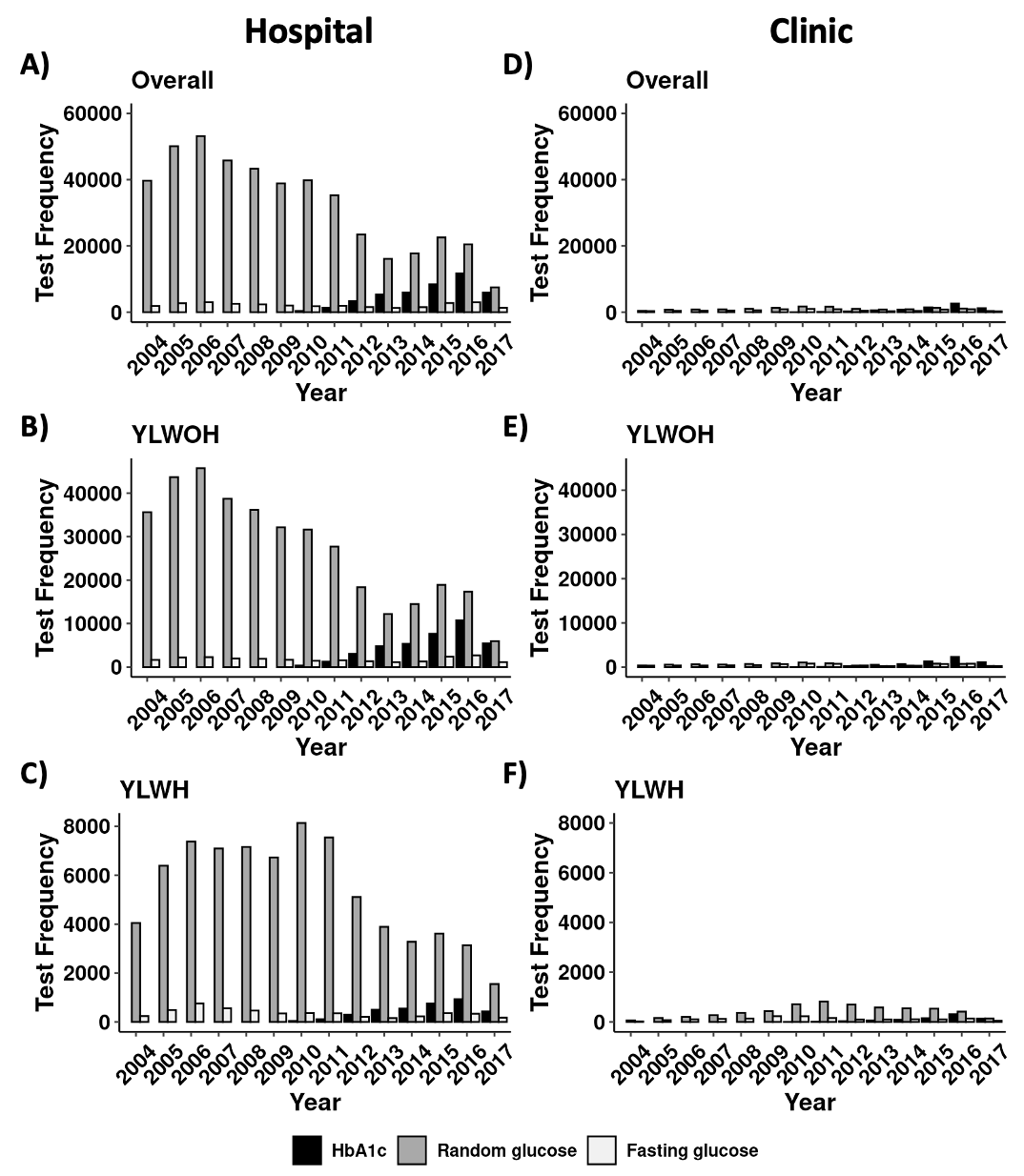

*YLWH – youth living with HIV; YLWOH youth living without HIV
