## Supplemental Table 1 for "Gaps in the Type 1 Diabetes Mellitus care cascade: a national perspective using South Africa’s National Health Laboratory Service (NHLS) database"

**Supplemental Table 1. Age at diagnosis for those with lab-diagnosed diabetes (N=256,449).**

|  |  |  | **For those with lab-diagnosed diabetes, age diagnosis was made** | | | | |
| --- | --- | --- | --- | --- | --- | --- | --- |
| **Province** | **Number screened** | **n (%) of lab**  **diagnosed diabetes** | **Overall** | **YLWOH**  **(n; median (IQR))** | **YLWH**  **(n; median (IQR))** | **Hospital**  **(n; median (IQR))** | **Clinic**  **(n; median (IQR))** |
| **Gauteng** | 111,631 | 13,264 (11.9%) | 13,264; 0 (0, 9) | 12,211; 0 (0, 9) | 1,053; 0 (0, 4) | 12,936; 0 (0, 8) | 328; 10 (0, 18) |
| **Kwa-Zulu Natal** | 14,758 | 2,159 (14.6%) | 2,159; 12 (5, 16) | 2,067; 12 (5, 16) | 92; 14 (7, 18) | 1,891; 12 (6, 16) | 268; 8 (0, 16) |
| **Western Cape** | 19,584 | 2,264 (11.6%) | 2,264; 9 (1, 14) | 2,187; 9 (1, 15) | 77; 1 (0, 5) | 2,173; 9 (1, 14) | 91; 16 (12, 18) |
| **Eastern Cape** | 26,232 | 3,339 (12.7%) | 3,339; 4 (0, 14) | 3,162; 4 (0, 14) | 177; 1 (0, 13) | 3,135; 3 (0, 14) | 204; 15 (5, 18) |
| **Free State** | 31,891 | 2,874 (9%) | 2,874; 1 (0, 12) | 2,727; 1 (0, 12) | 147; 3 (0, 14) | 2,793; 1 (0, 12) | 81; 15 (7, 17) |
| **Limpopo** | 19,421 | 2,878 (14.8%) | 2,878; 9 (1, 15) | 2,780; 9 (1, 15) | 98; 2 (1, 10) | 2,816; 9 (1, 15) | 62; 12 (0, 17) |
| **Mpumalanga** | 9,969 | 1,713 (17.2%) | 1,713; 11 (1, 16) | 1,593; 11 (1, 16) | 120; 6 (1, 15) | 1,540; 10 (1, 16) | 173; 11 (2, 17) |
| **Northern cape** | 2,501 | 313 (12.5%) | 313; 10 (1, 16) | 297; 11 (1, 16) | 16; 2 (0, 11) | 268; 10 (1, 15) | 45; 16 (3, 18) |
| **North West** | 20,462 | 2,206 (10.8%) | 2,206; 2 (0, 12) | 1,997; 2 (0, 13) | 209; 1 (0, 6) | 2,037; 1 (0, 12) | 169; 11 (0, 17) |
| **Total** | 256,449 | 31,010 (12.1%) | 31,010; 2 (0, 13) | 29,021; 2 (0, 13) | 1,989; 1 (0, 9) | 29,589; 2 (0, 13) | 1,421; 12 (1, 17) |

*YLWH – youth living with HIV; YLWOH youth living without HIV
